# Intraoperative superb microvascular ultrasound imaging in glioma: novel quantitative analysis correlates with tumour grade

**DOI:** 10.1101/2024.12.07.24318636

**Authors:** Luke Dixon, Alistair Weld, Dolin Bhagawati, Neekhil Patel, Stamatia Giannarou, Matthew Grech-Sollars, Adrian Lim, Sophie Camp

## Abstract

Accurate grading of gliomas is critical to guide therapy and predict prognosis. The presence of microvascular proliferation is a hallmark feature of high grade gliomas which traditionally requires targeted surgical biopsy of representative tissue. Superb microvascular imaging (SMI) is a novel high resolution Doppler ultrasound technique which can uniquely define the microvascular architecture of whole tumours. We examined both qualitative and quantitative vascular features of gliomas captured with SMI, analysing flow signal density, vessel number, branching points, curvature, vessel angle deviation, fractal dimension, and entropy. Results indicate that high-grade gliomas exhibit significantly greater vascular complexity and disorganisation, with increased fractal dimension and entropy, correlating with known histopathological markers of aggressive angiogenesis. The integrated ROC model achieved high accuracy (AUC = 0.95), highlighting SMI’s potential as a non-invasive diagnostic and prognostic tool. While further validation with larger datasets is required, this study opens avenues for SMI in glioma management, supporting intraoperative decision-making and informing future prognosis.

## Introduction

Adult-type diffuse gliomas are a group of primary infiltrative brain tumours which form the bulk of neuro-oncology practice. Whilst adult gliomas are relatively rare, they account for a disproportionate burden on society due to high mortality, representing the second leading cause of cancer-related death in those 15-39 years of age^1^. Prognosis varies widely depending on certain molecular and structural features. The recent 2021 WHO classification defines three types of adult-type diffuse glioma: glioblastoma *IDH*-wildtype (grade 4), astrocytoma isocitrate dehydrogenase (*IDH*) mutant (graded 2 to 4) and oligodendroglioma *IDH* mutant and *1p/19q* co-deleted (graded 2 or 3)^2,3^. Unfortunately, glioblastoma is the most common adult glioma and has the poorest prognosis, with a median survival of only 12-18 months. Whilst IDH mutant astrocytoma median survival rate ranges between 5-10 years (for grades 2 and 3) and 3-4 years (for grade 4), and oligodendroglioma survival rate is often above 10 years^4^. Discriminating the type and grade of glioma is imperative to guide treatment and predict prognosis. Although, in recent years, molecular features have taken a pre-eminent role in further stratifying gliomas, the presence of microvascular proliferation remains a pathological cornerstone for discriminating high-grade gliomas (HGG) from low-grade gliomas (LGG). Aberrant blood vessel growth is a defining feature of the highest grade, namely glioblastomas and grade 4 IDH mutant astrocytomas, and is characterised by multilayered small-calibre blood vessels.^5^ Microvascular proliferation is driven by several mechanisms, including peri-necrotic hypoxia promoted vascular epidermal growth factor expression.^6^ Presently, the detection of microvascular proliferation is mainly limited to histopathological assessment of biopsied tissue. This requires the sampled tissue to be representative, with a risk of underestimating grade if an area of tumour without microvascular proliferation is biopsied.

Advanced ultrasound (US) microvascular Doppler techniques, such as superb microvascular imaging (SMI), have recently been developed and translated into clinical practice. SMI is an emerging contrast-free, Doppler ultrasound technique developed by Canon Medical Systems (Tokyo, Japan), which uses an adaptive algorithm to filter out motion artefacts from true blood flow signals. This allows the delineation of a broader range of blood flow signals at a much higher resolution^7^. SMI has two main modes of operation: Color SMI (cSMI), which displays a standard grey-scale B-mode image with a colour-encoded Doppler signal and monochrome SMI (mSMI), which provides a grey-scale image of the vasculature only with greater sensitivity to blood flow. Outside of the CNS, SMI has been shown to improve the discrimination of malignant tumors from benign in several organ systems, including the breast and thyroid^8,9^. This is based on increased sensitivity to neovascularity, a common feature of many types of malignant tumors. There has been limited exploration of SMI and other similar advanced Doppler techniques in intraoperative brain tumor imaging^10–12^. Analysis of SMI images has also been mainly limited to qualitative evaluation of vascular morphology and measures of vessel density, as quantification of vascular shape and complexity is challenging^8,10,12^. This challenge is well recognized in other fields of microvascular imaging. In retinal microvascular photographs the application of novel mathematical measures of image complexity such as fractal dimension and entropy analysis plus automated techniques for measuring vascular branching points and curvature have been well established in differentiating and grading different pathological processes^13–15^. Recently some of these metrics have been translated and explored in a different type of high resolution microvascular doppler imaging developed in a research setting on other more accessible tissues such as the breast and choroid of the eye^16,17^.

In this exploratory study, we will use already clinically available SMI to generate a unique, intraoperative, whole tumour view of glioma microvascular architecture. We will then qualitatively assess and apply several novel quantitative methods already established in the analysis of retinal vessels and emerging in microvascular Doppler techniques to these SMI glioma images. Through this approach, we hope to explore whether SMI can detect microvascular proliferation and, in turn, discriminate high grade gliomas from low grade gliomas.

## Materials and Methods

### Ethics approval

The study had local ethical approval from Health Research Authority (HRA) and Health and Care Research Wales (HCRW) authorities (REC reference: 22/WA/0259). Patients were retrospectively recruited and the need for informed consent was waived. The study was performed in accordance with the relevant guidelines/regulations and performed in accordance with the Declaration of Helsinki.

### Study participants and inclusion and exclusion criteria

Patients with histologically confirmed adult-type diffuse glioma who underwent intraoperative ultrasound (ioUS) guided resection of their tumour at the Department of Neurosurgery, Charing Cross Hospital, Imperial College Healthcare NHS Trust (London, United Kingdom) were retrospectively included in this study between January 2020 and October 2024.

The inclusion criteria were: age >18 years; confirmed adult-type diffuse glioma based on histology; and the use of ioUS with SMI acquisitions. The exclusion criteria were: age <18 years; and absent or incomplete intraoperative US data.

### Intraoperative ultrasound protocol and SMI acquisition

Intraoperative ultrasound was performed with a latest generation Canon i900 Aplio US system with a high-frequency iDMS Micro-Convex probe (i8MCX1, imaging frequency 8-8MHz, frame rate 18 - 25 frames per second), provided by Canon Medical Systems (Otawara, Japan). Image acquisition was performed by a neuroradiologist and neurosurgeon experienced with ioUS and a set scanning protocol previously described by the authors was followed^18^. After craniotomy, before and after opening the dura, 2D B-mode US and SMI clips sweeping through the entire tumour were obtained in two orthogonal planes following optimisation of focal zone, depth and gain. SMI acquisitions were performed in dual screen mode with B mode and SMI displayed side by side to allow accurate discrimination of intratumoral versus surrounding cerebral vessels. Monochrome SMI was used due to its greater sensitivity versus colour SMI and is routinely used in our practice to assist with tumour detection and to identify important vascular structures. Saved US images and video clips were anonymised on the scanner prior to transferring onto a workstation for offline processing.

### Qualitative image analysis

Two methodologies were used to qualitatively classify the patterns of microvascular architecture. A semiquantitative grading system was used to classify the variation of blood vessel size observed within the tumor into four distinct grades: grade 0, indicating the absence of blood flow signals; grade 1, calibre matches normal parenchyma; grade 2, uniformly dilated vessels relative to normal parenchyma ; and grade 3, heterogenously varied calibre. The morphological characteristics of the vessels were also categorized into four patterns based on prior experience and influenced by similar work in classifying liver lesions on doppler: a, sparse, straight unidirectional, penetrating flow signal; b, absent central signal with normal appearing parenchymal vessels displaced around ; c, absent central signal with an irregular, tortuous, rim of vessels flow projecting to the hypovascular core; d, intrinsic, multidirectional, irregular, tortuous flow signal^19–21^. Patterns a and b were classified as hypovascular supply, while patterns c and d was classified as hypervascular supply. Two neurosurgeons both with over 5 years experience in intraoperative ultrasound performed the rating.

### Tumour vessel mask segmentation

For quantitative analysis, SMI images were first converted to the Nearly Raw Raster Data format (Nrrd) and then reviewed on 3DSlicer [https://www.slicer.org/] where three representative SMI images of the tumours were manually chosen by a neuroradiologist (LD) blinded to final histology, with 10 years of experience in ultrasound and 5 years experience in neurosurgical ioUS. Images with the least artefacts (e.g., transient brain pulsation artefact) were selected. The intratumoral microvascular tree was then manually segmented by the same neuroradiologist to mask out surrounding normal cerebral and extra-cerebral vasculature using the side-by-side B-mode image and preoperative navigation MRI as references. Initial segmentations were performed on the B-mode imaging, which were then cloned and translated to overlay the SMI image using fixed coordinates. Once overlaid on the SMI image, the segmentation was refined to remove any overlaid graphical user interface elements burnt to the image by the ultrasound machine and constrained to the SMI window. Extracerebral vascular elements (e.g. sulcal vessels) were also excluded from the segmentation to limit the segmentation to purely intratumoral vessels.

### Preprocessing of SMI masks

The SMI masks underwent two separate image processing steps to quantify both line-based and pixel-level features: 1) a simple noise robust, pixel intensity threshold for binarisation of the grayscale image, to assess flow signal density, and 2) vessel line detection and isolation for measurement of vascular properties such as curvature and fractal dimension.

The threshold binarisation was performed using Otsu’s automatic global thresholding method, a widely accepted technique for optimal threshold determining, to separate an image into a foreground (ones) and background (zeros) class via pixel intensity histogram analysis^22^. The method systematically evaluates a set of possible thresholds for separating the histogram into two distinct distributions, and the one which best maximizes the between-class variance and minimizes the within-class variance is chosen. For this work, the foreground is used to represent the flow signal and the background the low signal.

For the second method, the vessel lines were detected using the ridge detection plugin in Fiji (an ImageJ-based image processing software)^23^. This uses a well-established algorithm to identify curvilinear structures^24^. The algorithm detects 2D line points by applying Gaussian kernels and scale-space analysis to line profile models, enabling the detection of asymmetrical bar-shaped lines. These line points are linked into continuous lines and corresponding edge points along the normal direction are identified to accurately determine the true line positions; which are used for vessel measurement in this work. The following parameters for ridge detection were found to achieve optimal delineation of vessels line width 9.0, high contrast 230, low contrast 87, sigma 3.1, lower threshold 0.51 and upper threshold 1.53. From the plugin, each line is represented by a unique id and set of coordinates, and all lines belonging to an SMI image can then be saved in a CSV file.

### Quantitative image processing and feature extraction

The SMI images and corresponding binary maps and detected ridges were given a class label of HGG or LGG. The evaluation was performed using Python with the OpenCV, SciPy, Scikit-learn and SKimage libraries. Visualisation of the data was created using matplotlib and seaborn libraries.

For the binarised images, only flow signal density was measured. Flow signal density measures the ratio of white pixels (flow signal) to black pixels (background) within the segmented SMI region.

In the analysis of the detected ridges, the following seven characteristics were extracted: number of vessels, vessel length, curvature, number of branching points, vessel angle deviation, fractal dimension analysis, and entropy calculation. Number of vessels counted the number of vessels in each image. Vessel length measured the mean length of all lines in each image. Branching point counts the number of lines with more than two connections. Curvature measured the extent to which the lines changes direction along their paths, which is then averaged to reflect the overall tortuosity of the lines - a higher curvature suggests a more angled and irregular line and a lower curvature implies a straighter, less variable, line. The vessel angle deviation reflects the angular standard deviation of the mean resultant length which is a measure of the concentration of angles around a mean direction. This is based on the angles in radians, it sums up the sine and cosine of each angle and then calculates the magnitude of the resultant vector and normalizes it by dividing by the number of angles. The angular standard deviation uses the mean resultant length to calculate the standard deviation in angular data. A higher mean resultant length means angles are more concentrated around the mean direction, leading to a lower standard deviation. The purpose of the curvature and vessel angle deviation metrics is to measure the irregularity and degree of variation in the trajectories of the intratumoral vessels as increased disorganisation and variation of vessel orientations is well recognised in HGGs^25^. Vessel angle deviation has not been explored in SMI but this and related approaches looking at variance in vessel orientation have been applied in the analysis of retinal vasculature imaging and in cerebral vasculature on histological sampling^26,27^.

To quantify the structural complexity and overall organisation of the microvascular architecture, the fractal dimension and entropy of the ridges were measured. In fractal dimension analysis a box-counting method was used^28^. This approach divides the image into progressively smaller boxes and counts the number of boxes containing at least one white pixel. The slope of the logarithmic plot of the size of the box versus the count of the box approximates the fractal dimension, serving as a marker of structural complexity. The use of this novel quantification technique has been well explored in the analysis of other types of vascular imaging, such as in retinal vessel imaging, and has recently shown promise in the evaluation of similar microvascular Doppler imaging in other body systems such as the breast^13,17^. Entropy calculation was performed using Shannon entropy, which is a mathematical measure of uncertainty or randomness in a system. In image analysis, it quantifies how complex the pixel intensities are in an image, assessing how varied or uniform the image is. High entropy suggests a more complex distribution of structures within an image whilst low entropy reflects a more uniform or predictable structure. Measures of entropy have been explored in other medical and biological imaging analyses, but their utility in assessing microvascular imaging is underexplored^29,30^. An average of the final eight metrics across the three SMI images was then recorded for each glioma case.

### Statistical analysis

To assess the significance of each extracted microvascular metric for the HGG and LGG groups, normality was first tested using the Shapiro-Wilk test. Metrics that were normally distributed (p > 0.05) were evaluated with independent t-tests, while non-normally distributed metrics were analysed with the Mann-Whitney U test. The Pearson correlation matrix is then calculated to measure the metric interrelationships.

The potential of each metric for separating HGG and LGG was evaluated via receiver operating characteristic (ROC) analysis, with AUC values, optimal thresholds, and accuracies recorded. Then a linear regression model combined all metrics for an integrated ROC assessment of discriminative power.

Further analysis of the separability of the classes, using the investigated metrics, is conducted using Principal Component Analysis (PCA). To investigate the potential separability in PCA space, which is a popular technique for preparing data for machine learning algorithms such as support vector machines or K-nearest neighbour.

## Results

### Population and clinical characteristics

A total of 32 patients (13 females and 19 males) who underwent ioUS guided resection of brain tumours at our hospital were retrospectively recruited in the study, 22 patients with HGG and 10 with LGG. The pathological diagnosis according to the current WHO 2021 classification in the HGG group was 17 grade 4 IDH wild type glioblastoma, 3 grade 4 IDH mutant astrocytomas, 1 grade 3 anaplastic IDH mutant astrocytoma and 1 grade 3 anaplastic oligodendroglioma whilst in the LGG group there was 7 grade 2 IDH mutant astrocytomas and 3 grade 2 oligodendrogliomas (clinical information summarised in Table 1). Microvascular proliferation was reported in 15 of the 17 glioblastomas (88.2%), 3 of the grade 4 IDH mutant astrocytomas (75%) and in the sole grade 3 oligodendroglioma. Characteristically no microvascular proliferation was noted in the grade 3 IDH mutant astrocytoma or the low grade glioma group.

**Table 1.** Patient characteristics and tumour details for low grade gliomas (LGG) and high grade gliomas (HGG)

| Characteristics | LGG | HGG |
| --- | --- | --- |
| Total | 10 | 22 |
| Mean Age | 37.67 ± 10.06 | 55.31 ± 14.6 |
| Sex (N, %) |  |  |
| Female | 4 (12.5%) | 9 (28%) |
| Male | 6 (18.7%) | 13 (40.6%) |
| Tumour Location (N, %) |  |  |
| Frontal | 8 (25%) | 8 (25%) |
| Parietal | 1 (3.1%) | 6 (18.7%) |
| Temporal | 1 (3.1%) | 7 (21.9%) |
| Insula | - | 1 (2.9%) |
| Tumour type |  |  |
| IDH mutant Astrocytoma | 7 (Grade 2) | 4 (1 Grade 3, 3 Grade 4) |
| Oligodendroglioma | 3 (Grade 2) | 1 (Grade 3) |
| Glioblastoma IDH wild type | - | 17 (Grade 4) |
| Microvascular Proliferation | 0 | 18 (81.8%) |

### Qualitative features

The microvascular architecture was qualitatively different between the HGG and LGG groups in both vessel size and morphology (see Figure 2 for example images of different gliomas and Table 2 summarizing the qualitiative scoring). In vessel size the majority of LGGs were classified as having either normal or uniformly enlarged vessels (8/10, 80%) whilst in the HGGs the majority were classified as heterogenously variable in size by both readers (18/22, 90%, 20/22, 90.9%). In vessel morphology the LGGs were predominantly classified as exhibiting pattern A) Linear, penetrating (9/10 90%, 5/10, 50%) whilst in HGGs the majority were classified as either pattern (C) Irregular around a hypovascular core (7/22, 31.8%, 12/22, 54.5%) or pattern (D) Intrinsic irregular, multidirectional (12/22, 54.5%, 10/22, 45.5%). When grouping vessel size 1 and 2 as organised and 3 as disorganised there was a significant difference between both LGG and HGG groups and similarly when classifying morphology patterns (A) and (B) as hypovascular and (C) and (D) as hypervascular there was a significant difference between HGGs and LGGs. Cohen’s Kappa showed moderate agreement between both readers for both vessel size (0.476) and morphology pattern (0.422).

**Table 2.** Qualitative assessment of microvascular architecture by two readers with interobserver agreement and significance testing of features against final histological outcome of LGG or HGG. * Denotes a significant difference (p<0.05) in the metric between the two groups.

|  | LGG |  | HGG |  | Kappa | p-value |  |
| --- | --- | --- | --- | --- | --- | --- | --- |
|  | Reader 1 | Reader 2 | Reader 1 | Reader 2 |  | Reader 1 | Reader 2 |
| Vessel Size |  |  |  |  | 0.476 | **0.000\*** | **0.004\*** |
| 1. Normal | 5 | 2 | 1 | 0 |  |  |  |
| 2. Uniformly enlarged | 3 | 6 | 1 | 4 |  |  |  |
| 3. Variable | 2 | 2 | 20 | 18 |  |  |  |
| Vessel Morphology |  |  |  |  | 0.422 | **0.000\*** | **0.005\*** |
| A) Linear, penetrating | 9 | 5 | 1 | 1 |  |  |  |
| B) Absent with normal surrounding | 0 | 1 | 3 | 1 |  |  |  |
| C) Irregular around a hypovascular core | 0 | 3 | 7 | 11 |  |  |  |
| D) Intrinsic irregular, multidirectional | 1 | 0 | 12 | 10 |  |  |  |

**Figure 1.**
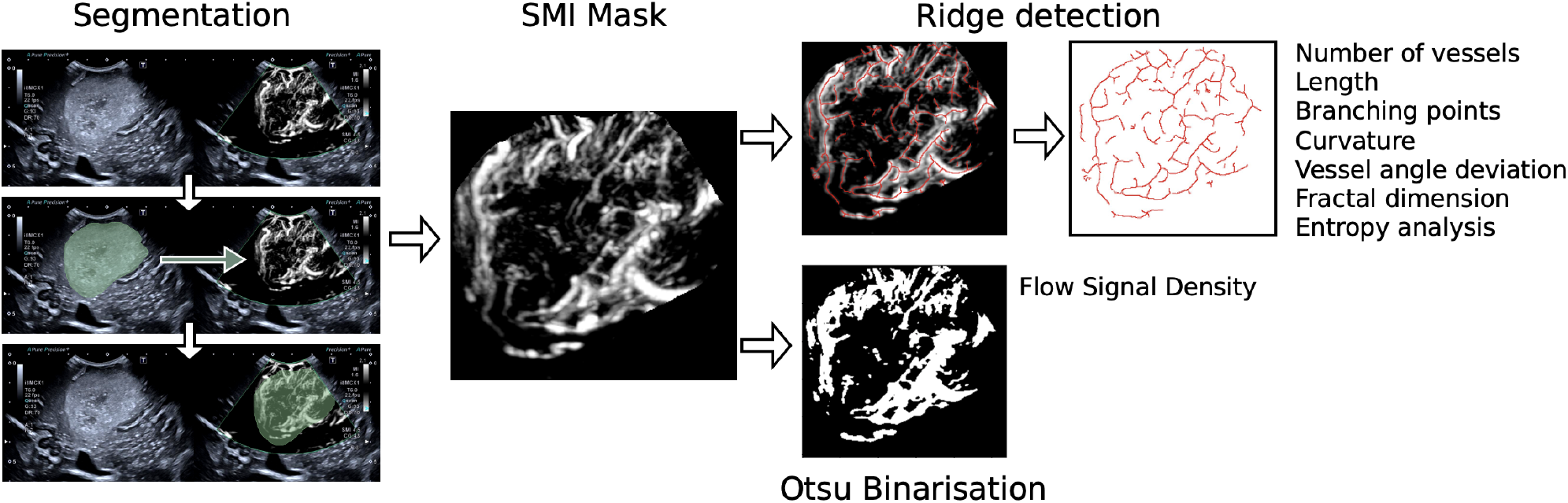
SMI Processing steps covering initial segmentation based on B-mode imaging on right then translated to mask the SMI image followed by ridge detection and Otsu binarisation of the masked SMI image.

**Figure 2.**
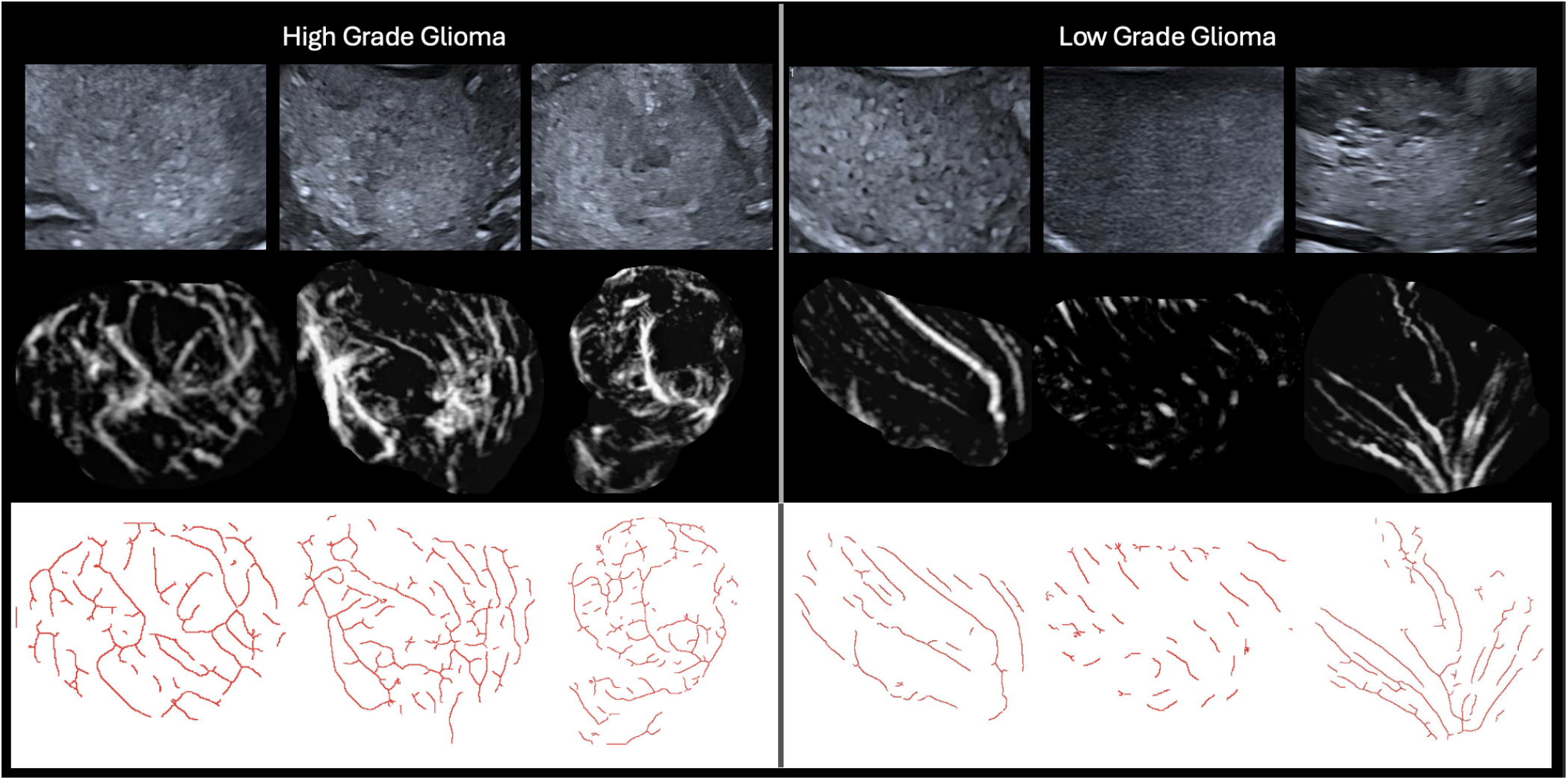
Examples of different HGGs (all IDH wild type grade 4 glioblastomas) and LGGs (all grade 2 IDH mutant astrocytomas) on B-mode (top row), SMI (middle row) and SMI following ridge detection (bottom row). Note the greater vascular density and more complex plus disorganised appearance to the microvascular architecture in the HGGs versus the more uniform and structured morphology of the vessels in the LGGs.

### Quantitative features

The results of the eight quantitative characteristics averaged between the HGG and LGG groups are summarised in Table 3. The distributions are visualised using the boxplots in Figure. 3. Seven of the eight metrics showed a significant difference between HGG and LGG. The flow signal density, number of vessels, branching points, curvature, vessel angle deviation, fractal dimension analysis, and entropy calculation were significantly higher in the HGG group while there was no significant difference in length. Fractal dimension analysis (AUC = 0.93, 95% CI: 0.87-0.98) and flow signal density (AUC = 0.92, 95% CI: 0.85-0.97) demonstrated the highest classification potential, achieving high accuracy at optimal thresholds (0.90 and 0.88, respectively) with significant p-values (p < 0.001). Branching points (AUC = 0.83 CI: 0.71-0.92) and number of vessels (AUC = 0.82 CI: 0.70-0.91) also showed a strong discriminative power, while entropy calculation, vessel angle deviation and curvature exhibited moderate classification capability, with AUC values of 0.77, 0.71, 0.68, respectively, and significant p-values (p < 0.01). Length displayed limited ability to distinguish HGG from LGG (AUC = 0.54, p = 0.535). A linear regression model integrating all features achieved the highest classification potential with an AUC of 0.95. Figure. 4 summarises the ROCs of the individual and pooled integated metrics and in Figure. -5 histograms of the distributions with the optimal threshold lines from the ROC analysis are presented.

**Table 3.** Comparison of various quantitative microvascular metrics between HGG and LGG groups. * Denotes a significant difference (p<0.05) in the metric between the two groups.

| Metric | HGG Mean (SD) | LGG Mean (SD) | p-value | AUC | 95% CI | Threshold | Accuracy |
| --- | --- | --- | --- | --- | --- | --- | --- |
| Flow Signal Density | 0.18 (0.09) | 0.06 (0.04) | **0.000\*** | 0.92 | (0.85 - 0.97) | 0.07 | 0.88 |
| Number of Vessels | 177.36 (71.87) | 100.37 (83.75) | **0.000\*** | 0.82 | (0.70 - 0.91) | 130.00 | 0.79 |
| Length | 158.41 (80.86) | 149.19 (86.95) | 0.535 | 0.54 | (0.41 - 0.67) | 121.20 | 0.60 |
| Branching Points | 64.09 (27.00) | 32.50 (30.38) | **0.000\*** | 0.83 | (0.71 - 0.92) | 39.00 | 0.81 |
| Curvature | 0.57 (0.03) | 0.55 (0.05) | **0.004\*** | 0.68 | (0.55 - 0.79) | 0.54 | 0.75 |
| Vessel Angle Deviation | 1.31 (0.14) | 1.18 (0.19) | **0.001\*** | 0.71 | (0.59 - 0.83) | 1.17 | 0.77 |
| Fractal Dimension | 1.04 (0.03) | 0.96 (0.03) | **0.000\*** | 0.93 | (0.87 - 0.98) | 1.00 | 0.90 |
| Entropy | 0.09 (0.03) | 0.06 (0.03) | **0.000\*** | 0.77 | (0.66 - 0.86) | 0.07 | 0.71 |
| Integrated | - | - | - | 0.95 | 0.91-0.99 | 0.63 | 0.92 |

**Figure 3.**
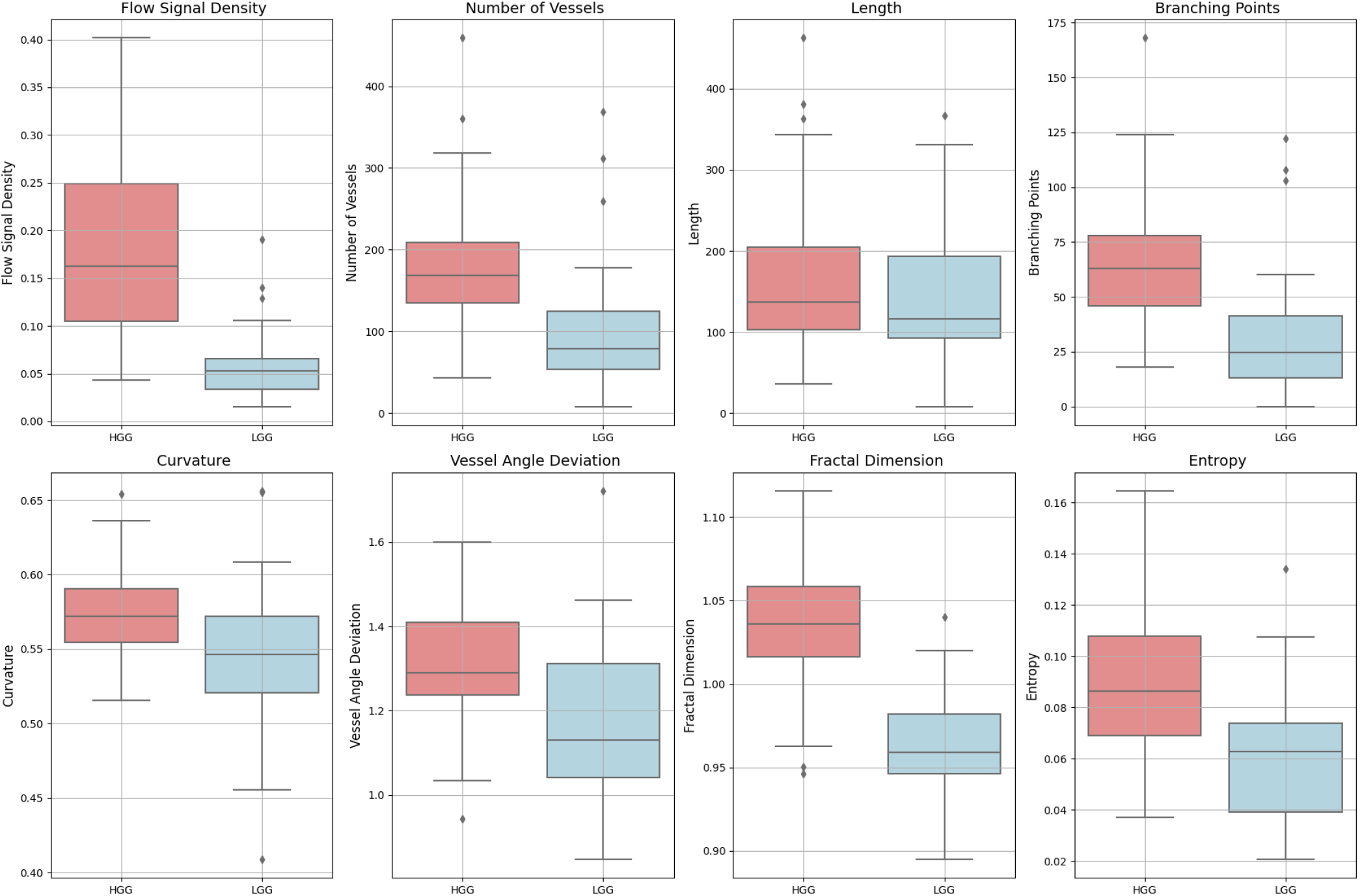
Boxplots of the distributions of the different quantitative microvascular metrics for HGG and LGG. Significant difference (p<0.05) between HGG and LGG was noted across all metrics except ‘Length”.

**Figure 4.**
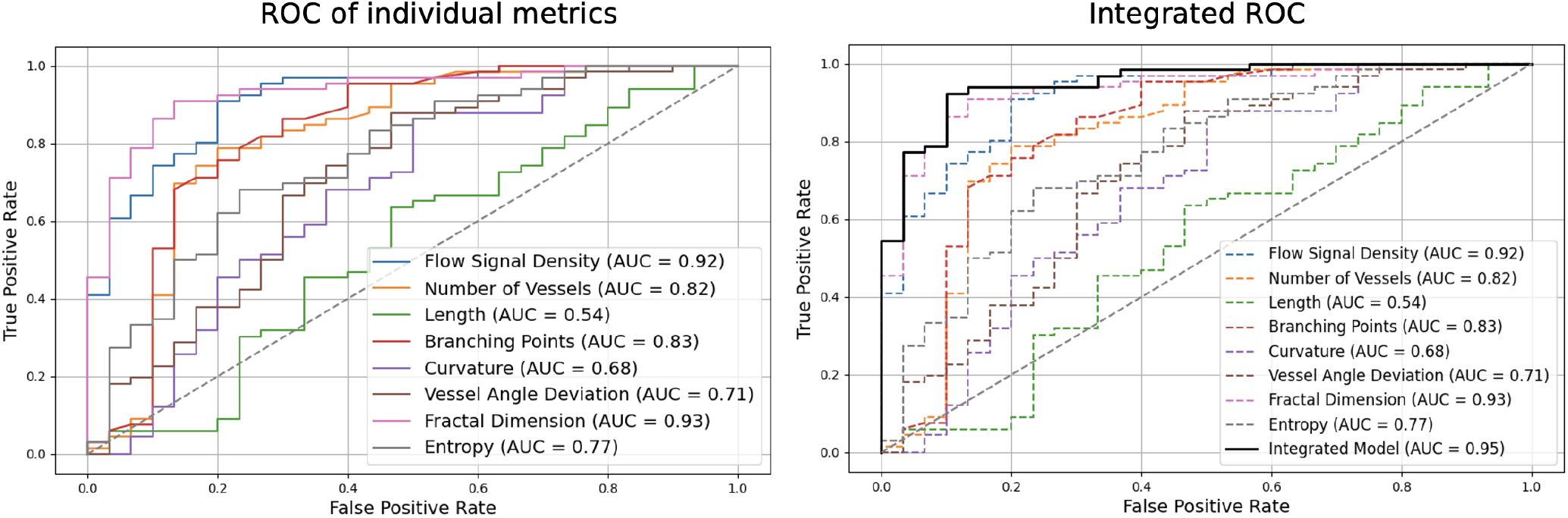
Summary of ROC of different quantitative microvascular metrics for differentiating HGG from LGG (right) plus integrated ROC of all metrics. Fractal dimension and flow signal density demonstrated the highest AUC whilst Length exhibited poor separation.

**Figure 5.**
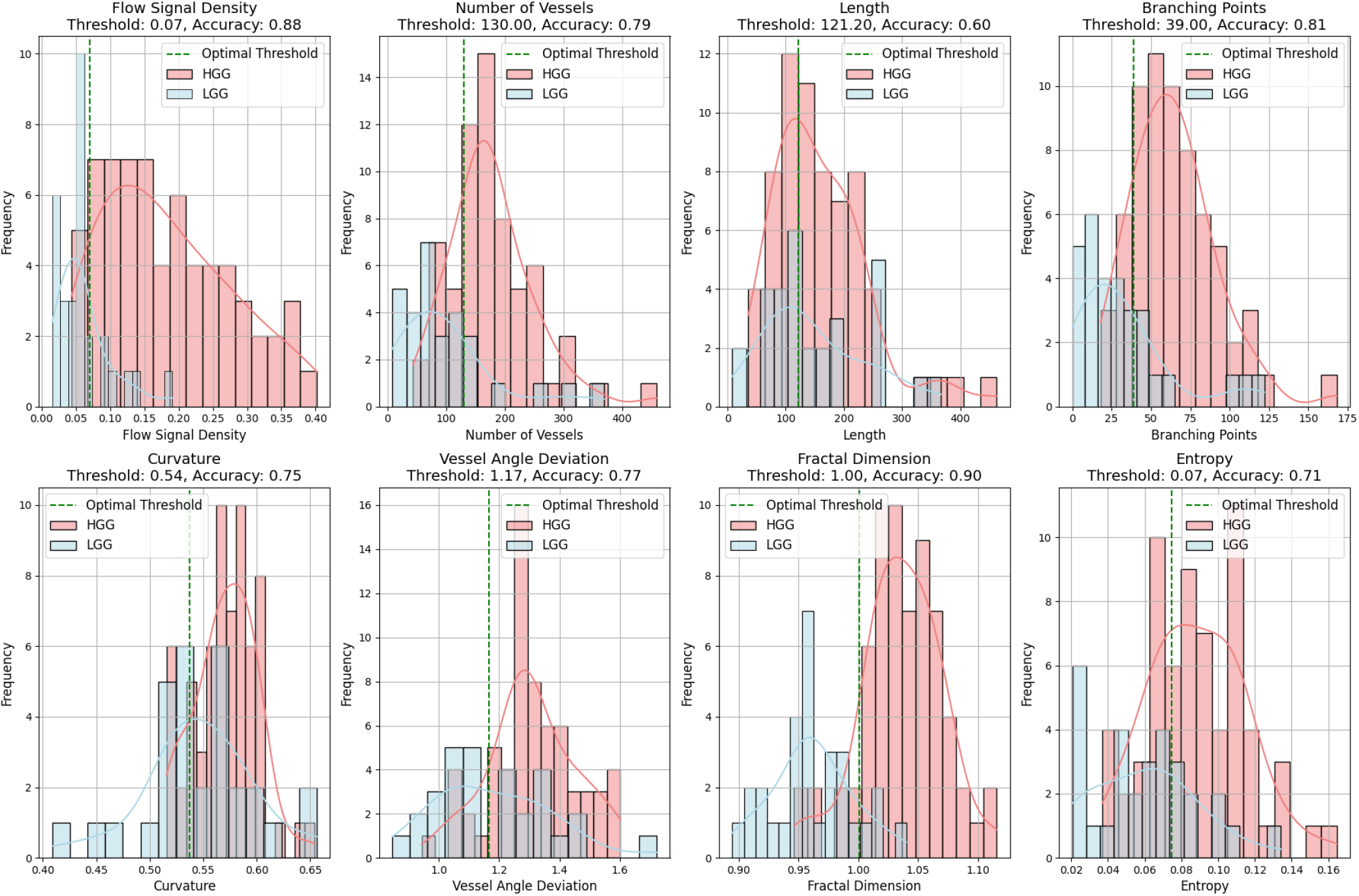
Histograms of the distributions of the different quantitative microvascular metrics for HGG and LGG with optimal threshold lines for discrimination derived from the ROC analysis.

A Pearson correlation matrix measuring the interrelationship of each microvascular metric is shown in Figure. 6. This found a very strong positive correlation with branching points and number of vessels (0.99). Strong correlations of entropy calculation with number of vessels (0.8) and branching points (0.8) plus fractal dimension analysis and flow signal density (0.77) and BP (0.66) were also observed. The length was weakly correlated with all other characteristics.

**Figure 6.**
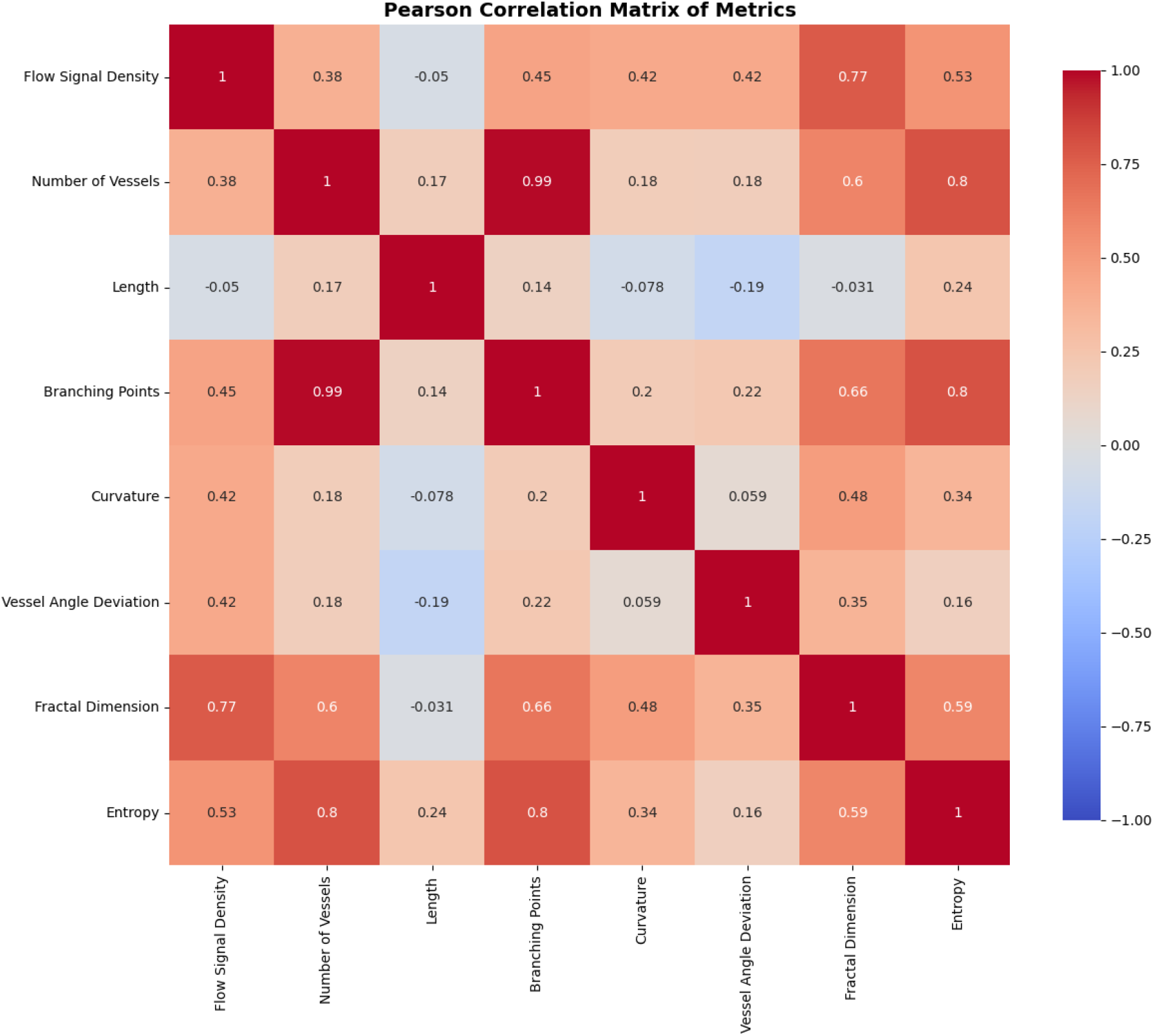
Pearson correlation matrix of microvascular quantitative metrics. Red cells reflect positive correlation and blue cells negative correlation. The majority of the metrics exhibited intermediate to moderation intercorrelation with only Branching points and Number of vessels showing almost equal correlation.

To complete the exploratory analysis and to further measure the separation of the microvascular metrics, the 2D PCA of the HGG and LGG groups is shown in Figure. 7. For visualisation, Kernel Density Estimation (KDE) is applied onto the 2D PCA plot, to show the overlap of the two groups. This visualisation suggests that there are shared microvascular features between the two classes, there is however appreciable clustering with the HGG group which appears more compact possibly reflecting more consistent features compared to the more dispersed LGG group. The explained variance for the two components is shown on the axes in the figure. PCA Component 1 explains 61.8% of the variance, and Component 2 explains 13.76%. Together, these two components account for 75.56% of the total variance, which is a strong representation of the data. This result suggests that the PCA projection and the first two components may be effective for machine learning classification of HGG and LGG.

**Figure 7.**
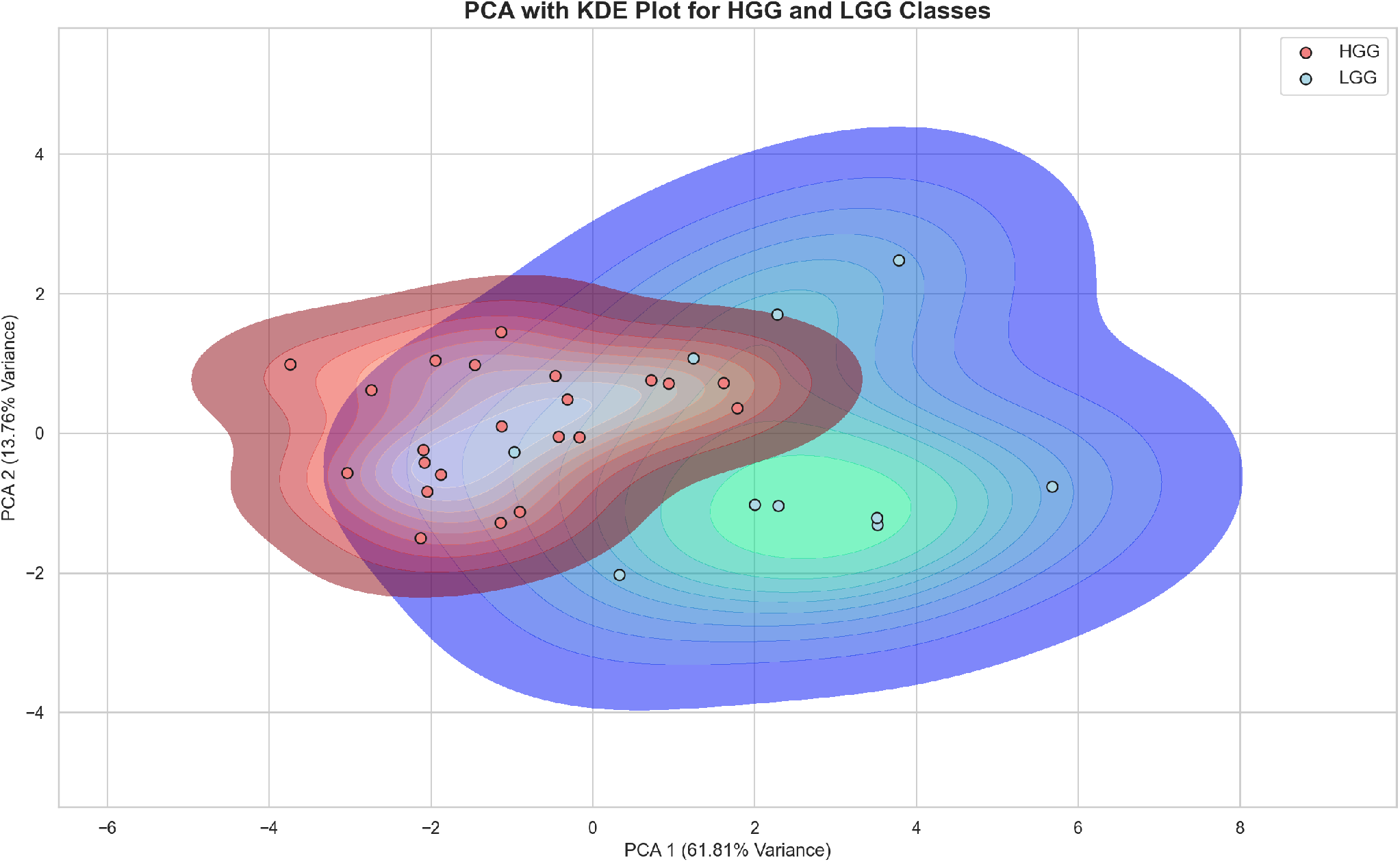
Principal component analysis of the microvascular metrics across the HGG and LGG groups with kernel density estimation (KDE) plot to visualise the underlying distribution of data across reduced dimensions. The plot shows overlap of the two classes but appreciable clustering of both groups suggesting that there are distinct features that with larger data sets may facilitate machine learning classification.

## Discussion

In this exploratory study, we have used an emerging, clinically available, non-contrast, high-resolution microvascular Doppler ultrasound technique called SMI to create a whole tumour view of the microvascular architecture in gliomas. Through applying adaptive algorithms to filter motion artifacts, SMI is able to clearly depict intratumoral microvasculature allowing more granular assesment of vascular morphology and density without the need for intravenous contrast agents. High-resolution microvascular doppler techniques like SMI offer a unique way of dynamically visualising the complex vascular network in brain tumours that other imaging modalities such as computerised tomography (CT) and magnetic resonance imaging (MRI) do not have the spatial or temporal resolution to resolve. The whole tumour view of vascularity is also generally not possible with histological sampling of gliomas, which, although of greater magnification, is typically only of small, discrete sections of the tumour, as opposed to the whole network. These tumours are typically removed in pieces and not en bloc like in other organ systems. Extending from this, we also showed that both qualitative scoring of microvascular characteristics and translation of a series of conventional plus more novel quantitative techniques, derived from other types of vascular analysis and emerging in high-resolution Doppler imaging, can help discriminate tumour grade^13,14,17^. To our knowledge, we are the first to apply these quantitative morphological parameters to microvascular Doppler imaging of gliomas. Of the measures, we found fractal dimension analysis, branching points, flow signal density and number of vessels offered the highest discriminatory power with moderate separation also noted with measures of vessel curvature, angulation and entropy. Our observation that flow signal density and vessel number correlate with HGGs parallels the histological observation of increased neoangiogenesis and microvascular proliferation in HGGs^25,31,32^. The vessel number metric showed a very high inter-metric correlation with the number branching points which was almost equal (0.99), suggesting that these effectively measure the same feature of vessel number. Interestingly, there was a weaker correlation between flow signal density and vessel number, implying that these capture discrete features, likely secondary to the flow signal density being influenced by the calibre of vessels as well as the number of vessels. Of all the metrics, fractal dimension analysis showed the highest AUC. Fractal dimension analysis is a measure of structural complexity and irregularity which quantifies how much detail is present in a structure at different scales. In the context of gliomas a high fractal dimension suggests a more complex and disordered vasculature which matches the pathological findings in HGG which are defined by the presence of heterogenous and irregular microvascular proliferation fueled by several factors including hypoxia-driven neoangiogenesis^25,31^. Furthermore, our observation of significantly higher curvature and vessel angle deviation in HGGs highlights the increased vessel tortuosity and disorganized orientation within these tumors. These metrics align well with the irregular, chaotic neoangiogenesis that characterizes HGGs, where rapid tumor expansion outstrips the formation of a structured blood supply, leading to a disordered vascular architecture^25,31,32^. This is further shown by the higher entropy feature seen in the HGG group which is a measure of randomness and disorder over the whole intratumoral microvascular image.

One of the clear limitations of SMI and other microvascular Doppler techniques in brain tumours is that imaging is not possible through the skull and is largely limited to the intraoperative setting where the tumour is already being resected or at least sampled. Despite this diagnostic limitation, these early insights into the potential unique role of quantitative microvascular Doppler imaging in brain tumour characterisation have several potential clinical implications that warrant further exploration. Firstly, whilst patients undergoing intraoperative SMI will inevitably have tissue sampling and definitive histology, tumour sampling in brain tumour surgery is often limited to small samples as the tumours are usually resected in parts or only biopsied particularly if there is involvement of an eloquent, highly functional part of the brain where the risk of morbidity or severe disability is considered too high. This can result in the sampled tissue not being representative of the most aggressive or highest-grade component or, at worst, not actually containing tumour. This risks underestimation of grade and the need for repeat surgery which has treatment implications. Considering this, our early work suggests that SMI and quantitative analysis may facilitate identifying the most high grade vascular components for resecting and sampling. This is supported by other recent preliminary work that demonstrated an improvement in the detection of brain tumor boundaries with SMI^33^. Secondly, prior histological studies have shown that there is a significant correlation between microvascular patterns and prognosis of HGGs. These patterns include microvascular sprouting, vascular clusters, vascular garlands, glomeruloid vascular proliferation and vascular mimicry, with the latter three patterns being associated with a poorer progression free survival and overall survival^34^. In light of this, it is possible that quantitative microvascular Doppler characterisation of microvascular patterns could serve as an additional prognostic biomarker that could be integrated into histology and molecular markers for predicting outcomes. In addition to this, there may also be a role in guiding therapy, for instance, different microvascular patterns may confer different responses to antiangiogenic therapies which inhibit abnormal vessel formation. This integrated multimodal approach would align with current trends in precision medicine and individualized glioma therapy. Finally, while limited to the intraoperative setting, there is early work showing that ultrasound monitoring of brain tumours is potentially feasible through sonolucent cranioplasty plates that can be placed at the time of surgical resection, which would further expand the utility of microvascular Doppler in the postoperative monitoring setting^35^.

This study has a few additional limitations. (i) While our initial exploratory analysis suggests that SMI and other high-resolution microvascular Doppler techniques may offer unique insights into glioma microvasculature, the sample size is limited and the generalizability of the tool and the applied quantitative methods is uncertain. (ii) There are also many potential variables in intraoperative US and SMI including operator experience, tumour location, tumour depth and artefacts which may impact the consistency of the quantitative metrics. Lastly, our study relies on manual segmentation to isolate the intratumoral vasculature, which is labour intensive and prone to subjective variability that will be influenced by operator experience.

Larger, potentially multi-site, studies are needed to further explore the discussed potentials of SMI and other microvascular Doppler techniques in brain tumour characterisation and treatment. Expanding the quantitative analysis across larger data sets and tumour types is also needed with correlation with clinial outcomes like progression free survival and overall survival. Although the sample size was small, our observation that the integrated metrics achieved a high AUC also suggests that with larger data sets a machine learning or deep learning approach to microvascular imaging may further refine microvascular imaging biomarkers.

## Conclusion

This exploratory study underscores the potential of SMI, an already clinically available imaging technique, as a new tool for glioma grading, providing real-time, dynamic imaging of the tumour microvascular architecture. To our knowledge, this is the first report of quantitative analysis of SMI metrics, such as fractal dimension analysis and flow signal density, which can provide an opportunity to define distinguishing features that may further refine the classification of glioma and help guide therapy. This research opens promising avenues for advancing brain tumour imaging.

## Data Availability

All data produced in the present study are available upon reasonable request to the authors.

## Declarations

The study had full local ethical approval by the Health Research Authority and Health and Care Research Wales (HCRW) authority. Study title - US-CNS: Multiparametric Advanced Ultrasound Imaging of the Central Nervous System Intraoperatively and Through Gaps in the Bone, IRAS project ID: 275556, Protocol number: 22CX7609, REC reference: 22/WA/0259, Sponsor: Research Governance and Integrity Team (RGIT). The study was performed in accordance with the relevant guidelines/regulations and performed in accordance with the Declaration of Helsinki.

## Data Availability Statement

The datasets used and/or analysed during the current study are available from the corresponding author on reasonable request.

## Acknowledgements

This research was supported by Canon Medical Systems, the NIHR Imperial Biomedical Research Centre, the UK Research and Innovation (UKRI) Centre for Doctoral Training in AI for Healthcare (EP/S023283/1), the Royal Society (URF \ R \ 2 01014]) and the Harmsworth Charitable Trust.

## Author contributions statement

L.D. conceived the experiments, collected the data, analysed the results and drafted the manuscript, A.W. contributed to the design of the experiments plus analysis, analysed the results and reviewed the manuscript, D.B. and N.P. contributed to the analysis as readers and reviewed the manuscript. A.L., S.G., M.GS. and S.C. reviewed the manuscript.

## References

1. Siegel, R. L., Miller, K. D., Wagle, N. S. & Jemal, A. Cancer statistics, 2023. CA: A Cancer J. for Clin. 73, 17–48, DOI: 10.3322/caac.21763 (2023). https://acsjournals.onlinelibrary.wiley.com/doi/pdf/10.3322/caac.21763.

2. Louis, D. N. et al. The 2021 WHO Classification of Tumors of the Central Nervous System: a summary. Neuro-Oncology 23, 1231–1251, DOI: 10.1093/neuonc/noab106 (2021).

3. McNamara, C. et al. 2021 WHO classification of tumours of the central nervous system: a review for the neuroradiologist. Neuroradiology 64, 1919–1950, DOI: 10.1007/s00234-022-03008-6 (2022).

4. Pekmezci, M. et al. Adult infiltrating gliomas with WHO 2016 integrated diagnosis: additional prognostic roles of ATRX and TERT. Acta Neuropathol. 133, 1001–1016, DOI: 10.1007/s00401-017-1690-1 (2017).

5. Nagashima, T., Hoshino, T. & Cho, K. G. Proliferative potential of vascular components in human glioblastoma multiforme. Acta Neuropathol. 73, 301–305, DOI: 10.1007/BF00686626 (1987).

6. Clara, C. A. et al. Angiogenesis and expression of PDGF-C, VEGF, CD105 and HIF-1α in human glioblastoma. Neuropathology 34, 343–352, DOI: 10.1111/neup.12111 (2014).

7. Fu, Z. et al. Clinical Applications of Superb Microvascular Imaging in the Superficial Tissues and Organs: A Systematic Review. Acad. Radiol. 28, 694–703, DOI: 10.1016/j.acra.2020.03.032 (2021).

8. Chen, L. et al. Additional Value of Superb Microvascular Imaging for Thyroid Nodule Classification with the Thyroid Imaging Reporting and Data System. Ultrasound Medicine & Biol. 45, 2040–2048, DOI: 10.1016/j.ultrasmedbio.2019.05.001 (2019).

9. Zhan, J., Diao, X.-H., Jin, J.-M., Chen, L. & Chen, Y. Superb Microvascular Imaging—A new vascular detecting ultrasonographic technique for avascular breast masses: A preliminary study. Eur. J. Radiol. 85, 915–921, DOI: 10.1016/j.ejrad.2015.12.011 (2016).

10. Ishikawa, M. et al. Ultrasonography Monitoring with Superb Microvascular Imaging Technique in Brain Tumor Surgery. World Neurosurg. 97, 11–749, DOI: 10.1016/j.wneu.2016.10.111 (2017).

11. Ishikawa, M. et al. Neurosurgical intraoperative ultrasonography using contrast enhanced superb microvascular imaging -vessel density and appearance time of the contrast agent-. Br. J. Neurosurg. 37, 485–494, DOI: 10.1080/02688697.2020.1772958 (2023).

12. Alafandi, A. et al. Probing the glioma microvasculature: a case series of the comparison between perfusion MRI and intraoperative high-frame-rate ultrafast Doppler ultrasound. Eur. Radiol. Exp. 8, 13, DOI: 10.1186/s41747-023-00406-0 (2024).

13. Doubal, F. et al. Fractal analysis of retinal vessels suggests that a distinct vasculopathy causes lacunar stroke. Neurology 74, 1102–1107, DOI: 10.1212/WNL.0b013e3181d7d8b4 (2010).

14. Cavallari, M. et al. Fractal Analysis Reveals Reduced Complexity of Retinal Vessels in CADASIL. PLoS ONE 6, e19150, DOI: 10.1371/journal.pone.0019150 (2011).

15. Yin, X., Ng, B. W.-H., He, J., Zhang, Y. & Abbott, D. Accurate Image Analysis of the Retina Using Hessian Matrix and Binarisation of Thresholded Entropy with Application of Texture Mapping. PLoS ONE 9, e95943, DOI: 10.1371/journal.pone.0095943 (2014).

16. Adusei, S. A. et al. Quantitative biomarkers derived from a novel, contrast-free ultrasound, high-definition microvessel imaging for differentiating choroidal tumors. Cancers 16, 395 (2024).

17. Ternifi, R. et al. Ultrasound high-definition microvasculature imaging with novel quantitative biomarkers improves breast cancer detection accuracy. Eur Radiol 32, 7448–7462, DOI: 10.1007/s00330-022-08815-2 (2022). 1432-1084 Ternifi, Redouane Wang, Yinong Gu, Juanjuan Polley, Eric C Carter, Jodi M Pruthi, Sandhya Boughey, Judy C Fazzio, Robert T Fatemi, Mostafa Alizad, Azra Orcid: 0000-0002-7658-1572 R01 CA239548/CA/NCI NIH HHS/United States R01CA168575/CA/NCI NIH HHS/United States P30 CA015083/CA/NCI NIH HHS/United States R01 CA168575/CA/NCI NIH HHS/United States R01 CA195527/CA/NCI NIH HHS/United States R01CA239548/CA/NCI NIH HHS/United States R01CA195527/CA/NCI NIH HHS/United States Journal Article Germany 2022/04/30 Eur Radiol. 2022 Nov;32(11):7448–7462. doi: 10.1007/s00330-022-08815-2. Epub 2022 Apr 29.

18. Dixon, L., Lim, A., Grech-Sollars, M., Nandi, D. & Camp, S. Intraoperative ultrasound in brain tumor surgery: A review and implementation guide. Neurosurg. Rev. 45, 2503–2515, DOI: 10.1007/s10143-022-01778-4 (2022).

19. Yang, F. et al. Superb microvascular imaging technique in depicting vascularity in focal liver lesions: more hypervascular supply patterns were depicted in hepatocellular carcinoma. Cancer Imaging 19, 92, DOI: 10.1186/s40644-019-0277-6 (2019).

20. Lee, D. H., Lee, J. Y. & Han, J. K. Superb microvascular imaging technology for ultrasound examinations: Initial experiences for hepatic tumors. Eur. J. Radiol. 85, 2090–2095, DOI: 10.1016/j.ejrad.2016.09.026 (2016).

21. He, M.-N., Lv, K.Jiang, Y.-X. & Jiang, T.-A. Application of superb microvascular imaging in focal liver lesions. World J. Gastroenterol. 23, 7765–7775, DOI: 10.3748/wjg.v23.i43.7765 (2017).

22. Otsu, N. A threshold selection method from gray-level histograms. IEEE Transactions on Syst. Man, Cybern. 9, 62–66, DOI: 10.1109/TSMC.1979.4310076 (1979).

23. Schindelin, J. E. et al. Fiji: an open-source platform for biological-image analysis. Nat. Methods 9, 676–682 (2012).

24. Steger, C. An unbiased detector of curvilinear structures. IEEE Transactions on Pattern Analysis Mach. Intell. 20, 113–125, DOI: 10.1109/34.659930 (1998).

25. Jain, R. K. et al. Angiogenesis in brain tumours. Nat. Rev. Neurosci. 8, 610–622, DOI: 10.1038/nrn2175 (2007).

26. Blinder, P. et al. The cortical angiome: an interconnected vascular network with noncolumnar patterns of blood flow. Nat Neurosci 16, 889–97, DOI: 10.1038/nn.3426 (2013). 1546-1726 Blinder, Pablo Tsai, Philbert S Kaufhold, John P Knutsen, Per M Suhl, Harry Kleinfeld, David DP1 OD006831/OD/NIH HHS/United States OD006831/OD/NIH HHS/United States R01 EB003832/EB/NIBIB NIH HHS/United States MH085499/MH/NIMH NIH HHS/United States R01 MH085499/MH/NIMH NIH HHS/United States R21 MH072570/MH/NIMH NIH HHS/United States MH072570/MH/NIMH NIH HHS/United States EB003832/EB/NIBIB NIH HHS/United States Journal Article Research Support, N.I.H., Extramural Research Support, Non-U.S. Gov’t United States 2013/06/12 Nat Neurosci. 2013 Jul;16(7):889–97. doi: 10.1038/nn.3426. Epub 2013 Jun 9.

27. Ma, Y., Ohr, M. P., Pan, X. & Roberts, C. J. Quantifying the pattern of retinal vascular orientation in diabetic retinopathy using optical coherence tomography angiography. Sci. Reports 11, 15826, DOI: 10.1038/s41598-021-95219-9 (2021).

28. Sarkar, N. & Chaudhuri, B. B. An efficient differential box-counting approach to compute fractal dimension of image. IEEE Trans. Syst. Man Cybern. Syst. 24, 115–120 (1994).

29. Xu, L. et al. Spatial distribution of the shannon entropy for mass spectrometry imaging. PLoS One 18, e0283966, DOI: 10.1371/journal.pone.0283966 (2023). 1932-6203 Xu, Lili Orcid: 0000-0001-5340-8841 Kikushima, Kenji Orcid: 0000-0001-8674-9554 Sato, Shumpei Islam, Ariful Sato, Tomohito Aramaki, Shuhei Zhang, Chi Sakamoto, Takumi Orcid: 0000-0003-2271-9975 Eto, Fumihiro Takahashi, Yutaka Yao, Ikuko Machida, Manabu Orcid: 0000-0002-1830-0760 Kahyo, Tomoaki Setou, Mitsutoshi Journal Article Research Support, Non-U.S. Gov’t United States 2023/04/07 PLoS One. 2023 Apr 6;18(4):e0283966. doi: 10.1371/journal.pone.0283966. eCollection 2023.

30. Karbalaeisadegh, Y., Yao, S., Zhu, Y., Grimal, Q. & Muller, M. Ultrasound characterization of cortical bone using shannon entropy. Ultrasound Med Biol 49, 1824–1829, DOI: 10.1016/j.ultrasmedbio.2023.04.006 (2023). 1879-291x Karbalaeisadegh, Yasamin Yao, Shanshan Zhu, Yong Grimal, Quentin Muller, Marie Journal Article England 2023/05/28 Ultrasound Med Biol. 2023 Aug;49(8):1824–1829. doi: 10.1016/j.ultrasmedbio.2023.04.006. Epub 2023 May 25.

31. Das, S. & Marsden, P. A. Angiogenesis in glioblastoma. N Engl J Med 369, 1561–3, DOI: 10.1056/NEJMcibr1309402 (2013). 1533-4406 Das, Sunit Marsden, Philip A P01 HL076540/HL/NHLBI NIH HHS/United States Journal Article United States 2013/10/18 N Engl J Med. 2013 Oct 17;369(16):1561–3. doi: 10.1056/NEJMcibr1309402.

32. Hardee, M. E. & Zagzag, D. Mechanisms of glioma-associated neovascularization. Am J Pathol 181, 1126–41, DOI: 10.1016/j.ajpath.2012.06.030 (2012). 1525-2191 Hardee, Matthew E Zagzag, David R21 NS074055/NS/NINDS NIH HHS/United States 1R21-NS074055-01A1/NS/NINDS NIH HHS/United States R01-CA100426-0141/CA/NCI NIH HHS/United States Journal Article Research Support, N.I.H., Extramural Research Support, Non-U.S. Gov’t Review United States 2012/08/04 Am J Pathol. 2012 Oct;181(4):1126–41. doi: 10.1016/j.ajpath.2012.06.030. Epub 2012 Aug 2.

33. Cai, S. et al. Clinical application of intraoperative ultrasound superb microvascular imaging in brain tumors resections: contributing to the achievement of total tumoral resection. BMC Med Imaging 24, 142, DOI: 10.1186/s12880-024-01321-5 (2024). 1471-2342 Cai, Siman Xing, Hao Wang, Yuekun Wang, Yu Ma, Wenbin Jiang, Yuxin Li, Jianchu Wang, Hongyan 2022-PUMCH-B-064/National High Level Hospital Clinical Research Funding./ Journal Article England 2024/06/12 BMC Med Imaging. 2024 Jun 11;24(1):142. doi: 10.1186/s12880-024-01321-5.

34. Chen, L. et al. Classification of microvascular patterns via cluster analysis reveals their prognostic significance in glioblastoma. Hum Pathol 46, 120–8, DOI: 10.1016/j.humpath.2014.10.002 (2015). 1532-8392 Chen, Long Lin, Zhi-Xiong Lin, Guo-Shi Zhou, Chang-Fu Chen, Yu-Peng Wang, Xing-Fu Zheng, Zong-Qing Journal Article Research Support, Non-U.S. Gov’t United States 2014/12/03 Hum Pathol. 2015 Jan;46(1):120–8. doi: 10.1016/j.humpath.2014.10.002. Epub 2014 Oct 14.

35. Del Bene, M. et al. Cranial sonolucent prosthesis: a window of opportunity for neuro-oncology (and neuro-surgery). J Neurooncol 156, 529–540, DOI: 10.1007/s11060-021-03929-x (2022). 1573-7373 Del Bene, Massimiliano Orcid: 0000-0002-6672-9721 Raspagliesi, Luca Carone, Giovanni Gaviani, Paola Silvani, Antonio Solbiati, Luigi Prada, Francesco DiMeco, Francesco Journal Article United States 2022/01/27 J Neurooncol. 2022 Feb;156(3):529–540. doi: 10.1007/s11060-021-03929-x. Epub 2022 Jan 26.

